# SARS-CoV-2 Vaccine Induced Atypical Immune Responses in Antibody Defects: everybody does their best

**DOI:** 10.1101/2021.06.24.21259130

**Authors:** Ane Fernandez Salinas, Eva Piano Mortari, Sara Terreri, Concetta Quintarelli, Federica Pulvirenti, Stefano Di Cecca, Marika Guercio, Cinzia Milito, Livia Bonanni, Stefania Auria, Laura Romaggioli, Giuseppina Cusano, Christian Albano, Salvatore Zaffina, Carlo Federico Perno, Giuseppe Spadaro, Franco Locatelli, Rita Carsetti, Isabella Quinti

## Abstract

**Background:** Patients with Primary Antibody Deficiencies (PAD) represent a potential at-risk group in the current COVID-19 pandemic. However, unexpectedly low cumulative incidence, low infection-fatality rate, and mild COVID-19 or asymptomatic SARS-CoV-2 infections were frequently reported in PAD. The discrepancy between clinical evidence and impaired antibody production requires in-depth studies on patients’ immune responses.

**Methods:** Forty-one patients with Common Variable Immune Deficiencies (CVID), 6 patients with X-linked Agammaglobulinemia (XLA), and 28 healthy age-matched controls (HD) were analyzed for anti-Spike and anti-RBD antibody production, generation of low and high affinity Spike-specific memory B-cells, Spike-specific T-cells before and one week after the second dose of BNT162b2 vaccine.

**Results:** HD produced antibodies, and generated memory B-cells with high affinity for Trimeric Spike. In CVID, the vaccine induced poor Spike-specific antibodies, and atypical B-cells with low affinity for Trimeric Spike, possibly by extra-follicular reactions or incomplete germinal center reactions. In HD, among Spike positive memory B-cells, we identified receptor-binding-domain-specific cells that were undetectable in CVID, indicating the incapability to generate this new specificity. Specific T-cell responses toward Spike-protein were evident in HD and defective in CVID. Due to the absence of B-cells, patients with XLA responded to immunization by specific T-cell responses only.

**Conclusions:** We present detailed data on early non-canonical immune responses in PAD to a vaccine against an antigen never encountered before by humans. From our data, we expect that after BNT162b2 immunization, XLA patients might be protected by specific T-cells, while CVID patients might not be protected by immunization.

## Introduction

The individual immune response to SARS-CoV-2 defines the COVID-19 clinical evolution, ranging from asymptomatic to mild, or moderate or severe disease with possible multi-organ failure requiring intensive care support [1].

Due the severely impaired immune response to infection and immunization, patients with Primary Antibody Deficiencies (PAD) [2] represent a potential at-risk group in the current COVID-19 pandemic [3]. However, an unexpected low number of SARS-CoV-2 infected patients have been reported [4-6] with a clinical presentation varying from mild symptoms to death, with many asymptomatic patients also documented. We have recently shown [7] that Italian PAD patients showed a cumulative incidence, and infection fatality rate similar to the SARS-CoV-2 positive Italian general population. It is possible to consider that the low incidence might be due the application of precautions measures our patients are used to follow since PAD diagnosis. The discrepancy between the clinical evidence and the defective antibody response underlines the limitations of this correlate of protection against the severe forms of COVID-19 and requires in-depth studies.

Vaccination is the safest and most effective tool to achieve a protective response in immunocompetent individuals in whom recent data demonstrated the high efficacy of SARS-CoV-2 immunization [8-9]. The European Society for Primary Immune Deficiency (ESID) recommends that PAD patients receive SARS-CoV-2 immunization provided that vaccines are based on killed/inactivated/viruses or on the use of mRNA [10]. The rationale is, as for the influenza immunization, that protection may be generated despite a low or even absent antibody response [11]. Data from PAD patients are needed, since it has been demonstrated that a weak selective pressure by an impaired immune response might fail to reach protection, increase the time of SARS-CoV-2 swab positivity [7], and also the risk of variants generation and spread [12]. In addition, the low or even absent antibody level is generating considerable anxiety in the PAD population aware of their incapacity to mount an adequate antibody response to infection and immunization [13]. We are running a study with the aim to define the short-and long-term mechanisms of impaired or preserved immune responses to SARS-CoV-2 immunization in a population of adult PAD patients. We present here data on early immune responses after BNT162b2 immunization. In a cohort of immunized PAD patients, we measured Spike-specific B- and T-cells and serum antibodies before immunization and one week after the second dose of the BNT162b2 vaccine. Results showed lack of antibody responses in two thirds of patients with Common Variable Immune Deficiencies (CVID), and in all patients with X-linked Agammaglobulinemia (XLA) who have no B cells but produce specific T-cell responses at the same extent of HD. CVID patients generated non-canonical B cell responses, as well as a variable response to the vaccination in terms of Spike-specific T-cells. This is the first study on the early B-cells response in PAD to a vaccine against an antigen never encountered before by humans.

## Methods

### Study Design and Patients

We studied patients regularly followed by the Italian Care Centers for adult patients with primary immune deficiencies in Rome and Naples. The study was carried out in 47 patients with PAD who agree to undergo SARS-CoV-2 immunization. We included also 28 immunized age-matched health care workers of the Bambino Gesù Children Hospital as healthy controls (HD). Eligible patients were informed on the study, including its safety profile and supply procedures, and signed the informed consents for vaccination and for the immunological study. The BNT162b2 vaccine was administered as prescribed, in two doses, 21 days apart. Two blood samples were obtained from each participant for serological and cellular immunity assessment at time 0 (T0), before the first dose, and seven days after the second dose (T1). During the study, patients were allowed to continue their therapies, and were monitored for their clinical status. The study was approved the Ethical Committee of the Sapienza University of Rome (Prot. 0521/2020, July 13, 2020). The study was performed in accordance with the Good Clinical Practice guidelines, the International Conference on Harmonization guidelines, and the most recent version of the Declaration of Helsinki. Methods for identify anti-Spike and anti-Receptor Binding Domain (RBD) antibodies and Spike-specific B- and T-cell responses are summarized in the Online Repository.

### Statistical analysis

Data were analyzed separately for the group of CVID patients, and for that of XLA patients and compared with data of age-matched controls (HD). Demographics were summarized with descriptive statistics (median and IQR for continuous values). Immunological, clinical variables were compared between the different study times. A univariate analysis assessed the impact of variable of interest. Values were compared by the non-parametric Kruskal-Wallis test and, if not significant, the Wilcoxon matched pair signed-rank test or the two-tailed Mann– Whitney U-test were used. Differences were deemed significant when P < 0.05. Statistical Package for Social Sciences version 15 (SPSS Inc., 233 South Wacker Drive, 11th Floor, Chicago) will be used for the analysis.

## Results

### Patients

In the study on Spike-specific antibody responses, we included 41 patients with CVID, 6 patients with XLA, and 28 healthy age-matched controls who received two doses of BNT162b2 vaccine. Demographic, clinical and immunological data at baseline of 47 patients enrolled and individual data are shown in Table S1. Immunoglobulin replacement treatment was continued in all PAD patients. Thirty-four CVID patients did not experience a previous SARS-CoV-2 infection, as shown by negativity at the periodical nasopharyngeal swab by PCR testing [7]. Seven CVID patients were vaccinated at least 3 months after recovering from a mild COVID-19. None PAD patients was infected with SARS-CoV-2 after completing the two doses vaccination, with the exception of one CVID patient who was infected two months after completing the vaccination, when his Spike-specific IgG level was 2.5 AU. He was treated with monoclonal antibodies within 24 hours from the onset of a mild fever and since then he did not show any additional COVID-19 symptoms. Twenty-six CVID, and 6 XLA patients, and 28 HD were included in the study of the specific B- and T-cell responses.

### SARS-CoV-2 antibodies

#### Common Variable Immunodeficiency

CVID is the most prevalent symptomatic PAD [14] with reduced or absent antibody response to infections and immunization, paucity of switched memory B-cells and dysregulated T-cell responses. Spike-specific IgG and IgA, and RBD-specific IgM were evaluated at T0 and T1. Antibodies to SARS-CoV-2 antigens in CVID patients and in HD are shown in Fig.1. In HD, anti-Spike IgG and IgA significantly increased in post-immunization samples, while anti-RBD antibodies of IgM isotype were already detectable at T0, reflecting the presence of natural or cross-reactive antibodies [15]. IgM increased at T1, with a wide variability between HD (Fig.1).

In 41 patients with CVID, we observed a significant increase in the median values of anti-Spike IgG (from T0: 0.12 AU (IQR 0.07-0.24) to T1: 0.39 AU (IQR 0.13-5.25), p<0.0001), but not of IgA (from T0: 0.05 AU (IQR 0.04-0.11) to T1: 0.07 (IQR 0.04-0.20), p=0.070). Anti-RBD IgM also increased (from T0: 0 µg/mL (IQR 0-0.42) to T1: 0.04 µg/mL (IQR 0.13-5.25), p=0.006) in CVID patients (Fig. 1). Post immunization antibodies against Spike and RBD were lower in CVID patients than in HD (IgG: p<0.0001, IgA: p<0.0001 and IgM: p=0.017) (Table 1). Post vaccination IgG highly correlated with post vaccination IgA (R: 0.8, p <0.0001) since 26 CVID patients did not respond, 4 patients responded with IgG and IgA, and 9 patients responded with IgG only. One patient responded with IgM only.

**Table 1.** Summary of antibody responses and specific B- and T-cells in CVID and XLA patients.

|  | HD<br>n=28 |  | HD<br>T0 vs T1 | CVID<br>n=26 |  | CVID<br>T0 vs T1 | CVID vs<br>HD (T1) | XLA<br>n=6 |  | XLA<br>T0 vs T1 | XLA vs HD<br>(T1) |
| --- | --- | --- | --- | --- | --- | --- | --- | --- | --- | --- | --- |
|  | Median | IQR (25th-75th) | P value | Median | IQR (25th-75th) | P value | P value | Median | IQR (25th-75th) | P value | P value |
| **SARS-Cov-2 antibodies** |  |  |  |  |  |  |  |  |  |  |  |
| **IgG AU, T0** | 0.10 | 0.00-0.15 | <0.0001 | 0.12 | 0.07-0.24 | <0.0001 | <0.0001 | 0.15 | 0.11-0.16 | >0.999 | <0.0001 |
| **IgG AU, T1** | 13.00 | 11-00-12.00 |  | 0.39 | 0.13-5.25 |  |  | 0.12 | 0.10-0.19 |  |  |
| **IgM µg/mL, T0** | 0 | 0-0.11 | <0.0001 | 0 | 0-0.42 | 0.006 | 0.017 | 0.00 | -- | -- | 0.004 |
| **IgM µg/mL, T1** | 1.5 | 1.5-3.0 |  | 0.04 | 0.13-5.25 |  |  | 0.00 | -- |  |  |
| **IgA AU, T0** | 0 | 0.2-0.3 | p< 0.0001 | 0.05 | 0.04-0.11 | 0.070 | <0.0001 | 0.13 | 0.05-0.22 | 0.562 | <0.0001 |
| **IgA AU, T1** | 9 | 11-13 |  | 0.07 | 0.04-0.20 |  |  | 0.05 | 0.04-0.14 |  |  |
| **Spike specific cells** |  |  |  |  |  |  |  |  |  |  |  |
| **MBC S+ %, T0** | 0.21 | 0.13-0.33 | 0.001 | 0.05 | 0-0.15 | 0.19 | <0.0001 | 0 | 0-0 | -- | <0.0001 |
| **MBC S+ %, T1** | 0.44 | 0.21-0.65 |  | 0.08 | 0-0.2 |  |  | 0 | 0-0 | -- |  |
| **MBC S++ %, T0** | 0 | 0-0.002 | <0.0001 | 0 | 0-0 | 0.06 | <0.0001 | 0 | 0-0 | -- | 0.0004 |
| **MBC S++ %, T1** | 0.016 | 0.005-0.04 |  | 0 | 0-0.002 |  |  | 0 | 0-0 | -- |  |
| **Activated MBC S+T0** | 0 | 0-0 | 0.06 | 0 | 0-0 | -- | <0.0001 | 0 | 0-0 | -- | <0.0001 |
| **Activated MBC S+ T1** | 0 | 0-0.46 |  | 0 | 0-0 |  |  | 0 | 0-0 | -- |  |
| **Activated MBC S++ T0** | 0.0 | 0-0 | 0.030 | 0 | 0-0 | -- | <0.0001 | 0 | 0-0 | -- | <0.0001 |
| **Activated MBC S++ T1** | 0.21 | 0-0.65 |  | 0 | 0-0 |  |  | 0 | 0-0 | -- |  |
| **ATM S+ %, T0** | 0 | 0-0 | 0.0001 | 0.0 | 0-0 | 0.019 | 0.43 | 0 | 0-0 | -- | 0.08 |
| **ATM S+ %, T1** | 0.06 | 0-0.35 |  | 0.0 | 0-0.31 |  |  | 0 | 0-0 | -- |  |
| **AMT S++ %, T0** | 0 | 0-0 | 0.250 | 0 | 0-0 | >0.999 | 0.4 | 0 | 0-0 | -- | >0.999 |
| **ATM S++ %, T1** | 0 | 0-0 |  | 0 | 0-0 |  |  | 0 | 0-0 | -- |  |
| **PB S+ %, T0** | 0 | 0-0 | <0.0001 | 0 | 0-0 | 0.001 | 0.027 | 0 | 0-0 | >0.999 | 0.0006 |
| **PB S+ %, T1** | 1.61 | 0.67-2.7 |  | 0.72 | 0-1.81 |  |  | 0 | 0-0.25 |  |  |
| **PB S++ %, T0** | 0 | 0-0 | <0.0001 | 0 | 0-0 | 0.125 | 0.005 | 0 | 0-0 | >0.999 | 0.17 |
| **PB S++ %, T1** | 0.15 | 0-0.51 |  | 0 | 0-0 |  |  | 0 | 0-0.25 |  |  |
| **RBD+ %, T0** | 1 | 0-3 | <0.0001 | 0-0 | 0-0 | -- | <0.0001 | 0-0 | 0-0 | -- | <0.0001 |
| **RBD+ %, T1** | 16 | 4-39.75 |  | 0-0 | 0-0 | -- |  | 0-0 | 0-0 | -- |  |
| **Spike-specific T cells**<br>**(IFN-g SFU x 10<sup>6</sup> cells) T0** | 2.5 | 0-68.7 | 0.041 | 0 | 0-9 | 0.151 | 0.001 | 37.5 | 0-50.5 | 0.041 | 0.937 |
| **Spike-specific T cells**<br>**(IFN-g SFU x 10<sup>6</sup> cells) T1** | 115 | 70-250 |  | 7 | 0-35 |  |  | 152.5 | 41.2-185 |  |  |

Abbreviations: ATM S+ Atypical Memory B-cells with low affinity for Trimeric Spike, ATM S++ Atypical Memory B-cells with high affinity for Trimeric Spike, CVID common variable immunodeficiency, HD healthy donors, IFN-g Interferon gamma; IQR interquartile range; MBC memory B-cells with low (S+) or high (S++) affinity for Trimeric Spike, PB Plasmablasts with low (S+) or high (S++) affinity for Trimeric Spike; RBD Receptor Binding Domain; XLA X-linked Agammaglobulinemia

**Figure 1.**
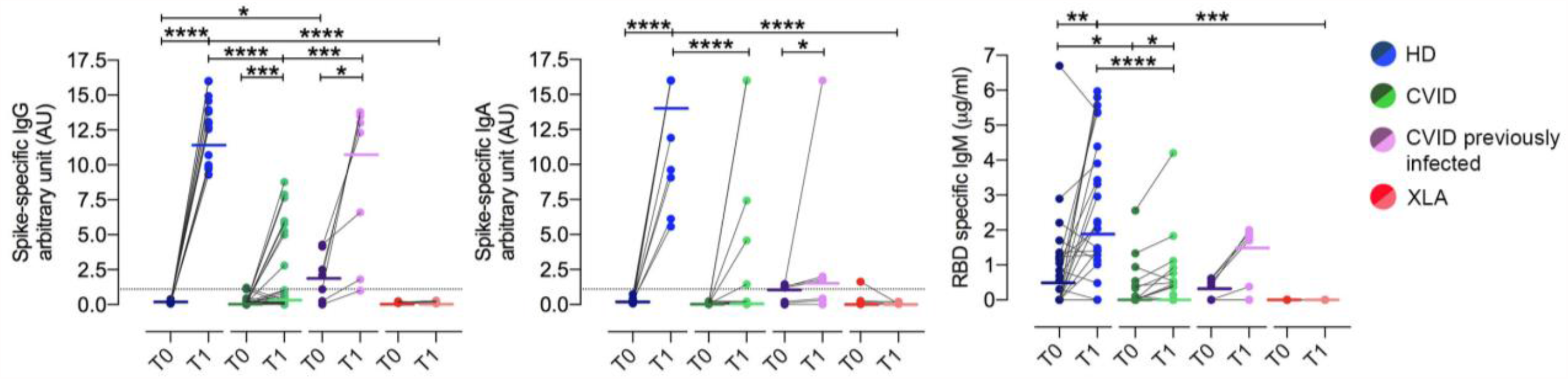
RBD-specific IgM and Spike-specific IgG, and IgA antibodies in HD (blue circles), CVID (green circles), CVID previously infected patients (pink circles), and XLA (red circles), before (T0, dark color circles) and one week after the second dose (T1, lighter color circles) of BNT162b2 vaccine. For each group the median is shown as a bar. * P ≤ 0.05, ** P ≤ 0.01, *** P≤ 0.001, **** P<0.0001. Positive cut-off value is represented by a black line. N = 28 HD, N= 34 CVID, N=6 XLA patients, and N=7 CVID previously SARS-Cov-2 infected.

IgG were already detectable at T0 (median 2.1AU (IQR 0.25-4.15) in five out of seven CVID patients who were previously infected with SARS-CoV-2 (3-8 months before vaccination), suggesting that IgG might persist after primary infection in some patients; in six out of seven patients, IgG increased at T1 (median 12.31 AU (IQR 1.8-13.50), showing that IgG were boosted by the subsequent immunization with two doses of BNT162b2 vaccine (Fig. 1).

#### X-linked Agammaglobulinemia

As expected, due to the lack of B cells and the consequent lack of serum antibody in XLA [16], anti-Spike and anti-RBD specific antibodies were not generated after immunization (Fig. 1).

### Spike SARS-CoV-2 memory B cells

High specificity and affinity are the most important characteristics of protective memory B-cells (MBCs), generated by the adaptive immune system in response to infection or vaccination [17]. MBCs, atypical memory B cells (ATM), activated MBCs, and plasmablasts (PBs) were identified by flow-cytometry, based on the expression of CD19, CD27, CD24 and CD38 markers (Fig. 2). MBCs were identified as CD19+ CD24+ CD27+ CD38-cells; ATMs were identified as CD19+ CD27-CD24-CD38-cells, that also are also CD21 negative [18,19]. Activated MBCs also lack CD21, but express CD27 [20,21]. PBs were identified as CD19+CD24-CD38++CD27++ cells. Overall, CVID patients showed lower frequency of MBCs, and higher frequency of ATMs and PBs than HD at all study times (Fig.3, and Table 1). This baseline pattern that distinguishes the B-cell populations of HD from those of CVID patients affects the specific response to vaccination.

**Figure 2.**
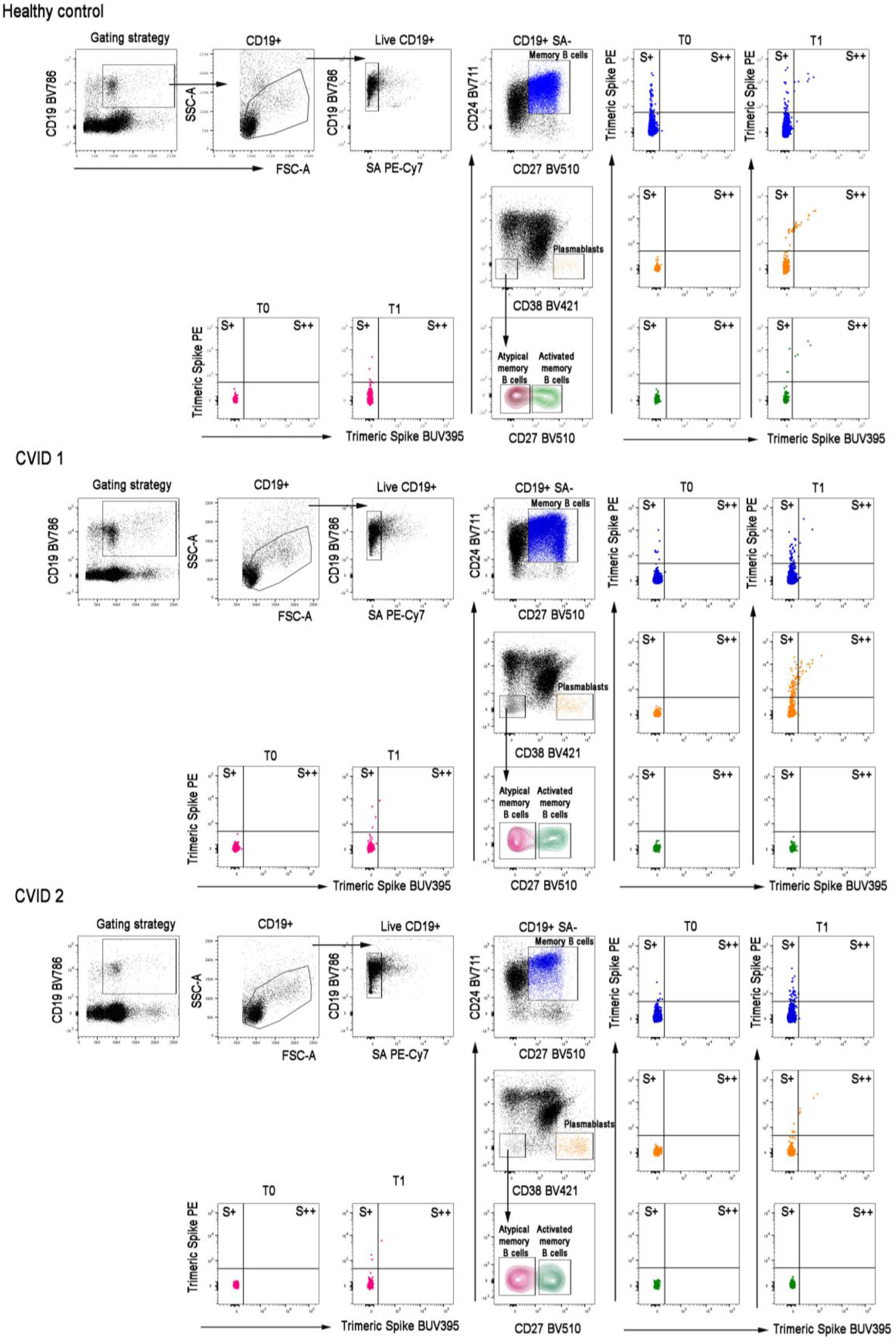
Gating strategy to identify S+ and S++ MBCs, PBs, ATM and activated MBCs. One HD and two CVID subjects are shown. We analyzed CD19+ B cells, included in live gate. SA PE-Cy7 was used as a decoy probe to gate out streptavidin binding B cells from further analysis. MBCs were identified as CD24+CD27+CD38-; PBs as CD24-CD27++CD38++; ATM as CD24-CD27-CD38-CD21- and activated MBCs as CD24-CD38-CD21-CD27+. Flow cytometry plots show the staining patterns of SARS-CoV-2 antigen probes in the indicated B cell populations during the follow-up. S+ are B cells that are Spike-PE+, but Spike-BUV395-. S++ are instead Spike-PE+ and SpikeBUV395+. The color code identifies Spike+ and ++ in MBC (blue), PBs (orange), ATM (dark red) and Activated MBCs (green).

B-cells specific for SARS-CoV-2 Spike protein were distinguished by their ability to bind biotin-labelled Trimeric Spike. Thanks to the availability of biotin-labelled Trimeric Spike coupled to an extremely high brightness fluorescence dye (PE) and the same biotin-labelled Trimeric Spike coupled to a moderate brightness fluorescent dye (BUV395) [22] (Fig.2, gating strategy) we were able to distinguish MBCs with low (PE single positive, S+) or high affinity (PE-BUV395 double positive, S++) for Trimeric Spike. Receptor binding domain (RBD) B cells were identified by RDB-biotin labeled with streptavidin-FITC.

All HD responded to immunization by generating a classical B cell response with low and high affinity MBCs (p=0.001 and p<0.0001, respectively) and high affinity activated MBCs (p=0.030). HD generated low affinity Trimeric Spike ATMs (p=0.0001), and low and high affinity Trimeric Spike PBs (p<0.0001) (Fig. 3 and Table 1), demonstrating that these populations are induced by the immune response to vaccination [23].

**Figure 3.**
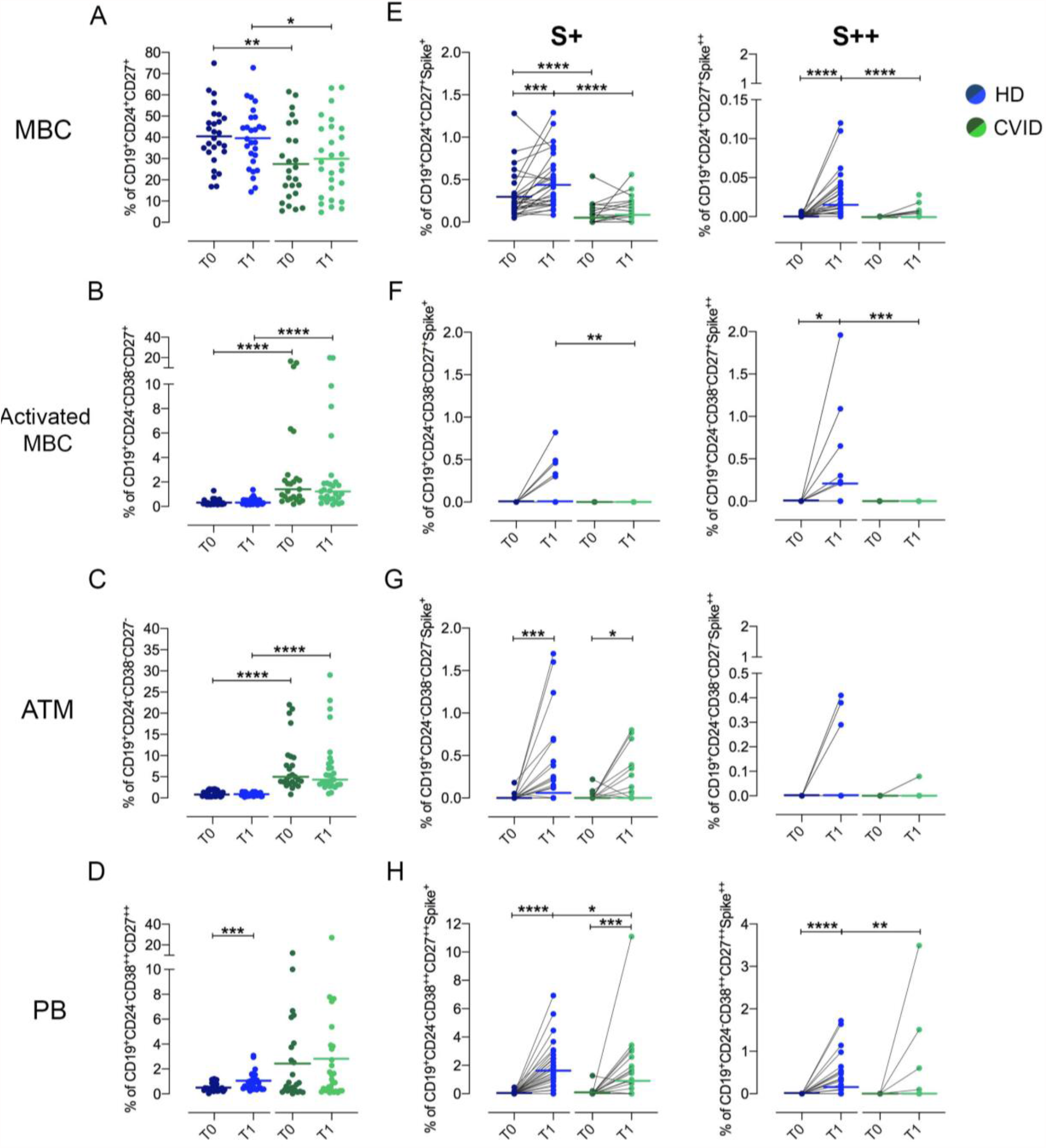
Peripheral blood B cells subsets, high and low affinity B cells for Trimeric Spike in HD (blue circles) and in CVID (green circles) before (T0, dark color circles) and after two doses of the BNT162b2 vaccine (T1, lighter color circles). We show the frequencies of peripheral blood MBCs (panel A), Activated MBCs (B), ATM (C) and PBs (E) in HD (blue circles) and CVID (green circles). The frequency of S+ and S++ B cells inside each identified B cell population is shown (E, F, G and H). Median are plotted as horizontal bars and statistical significances were determined using two-tailed Mann–Whitney U-test or Wilcoxon matched-pairs signed-rank test. *p < 0.05, **p < 0.01, ***p < 0.001; ****p< 0.0001. N = 28 HD and N= 32 CVID patients.

On the contrary, patients with CVID did not generate classical and activated MBCs, but vaccination induced Trimeric Spike ATMs with low affinity only (p=0.019) (Fig.3 and Table 1). This was observed only in about one third of CVID patients, where the frequency of low affinity ATMs at least doubled seven days after the second dose of vaccine. ATMs originally observed in tonsils and later in peripheral blood, are observed in conditions of chronic antigen stimulation [23] and may be produced by extra-follicular reactions or failed and incomplete germinal center reactions [24,25]. As expected, high affinity PBs for Trimeric Spike were produced in HD at T1, but not in CVID, with the exception of three patients who also produced specific IgG and IgA. Most CVID patients were able to generate low affinity Trimeric Spike PBs (p=0.001) that are possibly short-lived PBs derived from the non-canonical pathway of ATMs. Among total Spike positive MBCs (S+ plus S++), we also identified RBD-specific cells. RBD+ cells represent a minority of the MBCs generated by vaccination and significantly increased at T1 in HD (p<0.0001) (Table 1 and Figure S1). These cells, with high specificity against the virus RBD and thus producing most of the neutralizing antibodies, are undetectable in CVID patients indicating the incapability of CVID B-cells to undergo somatic mutation in the germinal center and thus generate this new specificity.

XLA patients lacks B cells and consequently did not generate any specific B-cells (Table 1).

### Spike SARS-CoV-2 T-cells

In HD, vaccination induced the generation of Spike-specific T-cells evaluated as Interferon gamma (IFN-g) Colony Forming Units that significantly increased (from T0: median 2.5 SFU (IQR 0-0.69) to T1: 115 median SFU (IQR 0.13-5.25), p=0.041). Differently from what observed after influenza vaccination [26], in CVID patients, Spike-specific T-cells producing IFN-g did not increase (from T0: median 0 SFU (IQR 0-9) to T1: median 7 SFU (IQR 0-35), p=0.151) and were significantly lower than in HD (p=0.0001). Nevertheless, XLA patients developed a significant T-cell response to vaccination. Indeed, Spike-specific T-cells were detected in 5 out of 6 tested patients (from T0: median 37.5 SFU (IQR 0-50.5) to T1: median 152.5 SFU (IQR 41-185), p=0.041), with a negligible difference respect to the one established in HD (p=0.937).

## Discussion

Effective vaccines against SARS-CoV-2 are being administered worldwide with the aim of terminating the COVID-19 pandemic. As for all immunizations, the efficacy has been linked to the production of specific antibodies, which increase in response to all vaccines in use [27,28]. The majority of patients with PAD show clinical and immunological characteristics implicating a functional impairment of B cell compartment, and a dysregulation of T-cell responses causing hypo-gammaglobulinemia or a-gammaglobulinemia and susceptibility to a wide range of microbial infections [29]. Despite severe impaired antibody responses, when infected with SARS-CoV-2, one fourth of adult PAD patients remained asymptomatic and half of them showed a mild disease [7]. It should be considered that a protective effect against severe COVID-19 could be due to the immune-modulatory effects on innate immunity exerted by immunoglobulin therapy also when administered at replacement dosages [30]. However, the discrepancy between this clinical evidence and the lack of antibodies underlines the limitations of this correlate of protection against the severe forms of COVID-19 and requires in depth studies on SARS-CoV-2 specific B- and T-cells function in PAD. However, data on immunogenicity of SARS-CoV-2 vaccine in patients with inborn errors of immunity are few and limited to anecdotal cases or heterogeneous cohort [31]. Although the natural course of COVID-19 is primarily characterized by the function of the innate immune system, with a secondary involvement of T- and B-cells, vaccines are designed to force the adaptive immune system to generate neutralizing antibodies and memory B- and T-cells that effectively protect from COVID-19.

Here we showed that while HD produced specific antibodies and generate high affinity MBCs and activated MBCs that significantly increased after immunization, these responses are lacking in all XLA and severely impaired in CVID patients after SARS-CoV-2 immunization, suggesting an incomplete protection. However, in five out of seven CVID patients who were previously infected with SARS-CoV-2, IgG were detectable at the time of vaccination administered at least three months after SARS-CoV-2 infection recovery, suggesting that if IgG were produced, they might persist after the primary infection. Moreover, in these previously infected CVID patients, IgG response was boosted by the subsequent immunization.

Interestingly, in all CVID patients, vaccination induced Trimeric Spike-specific B-cells inside the ATM and PB population, with low affinity for Trimeric Spike, possibly suggesting that the B-cell responses occurred mostly at extra-follicular sites [32] as recently demonstrated [33]. In line with this hypothesis, RBD+ B-cells were undetectable in CVID patients, whereas they represent 20% of the highly specific anti-Trimeric Spike response in HD (unpublished data) able to develop and successfully terminate the germinal center reaction [34]. Thus, CVID patients were able to respond to immunization by two doses of BNT162b2 with non-canonical lineage B-cells induced by a primary exposure to a novel antigen. It has been suggested that atypical B cells are short-lived activated cells, in the process of differentiating into plasmacells [23,35]. In addition, interesting information was gained by the parallel study of the T-cell responses, showing a robust generation of Spike-specific T-cells in all but one patient with XLA, and in HD. Specific-T-cell responses were induced in a minority of CVID patients with a variable frequency. Based on data on response to influenza virus immunization [11], we expected a more efficient generation of specific T-cells. However, this was not the case. While after influenza virus immunization T-cells are generated after multiple exposures to viral antigen following infection and immunization, SARS-CoV-2 is a pathogen never encountered before. Then, it is possible that the first antigenic stimulation was not sufficient to induce an early T-cell response.

Based on these data, how can we explain the paucity of symptoms or the mild COVID-19 course in PAD patients, and what might we expect after immunization?

To pathogens for which there is no preexisting immunity, our organism reacts by rapidly engaging the innate immune system with the intent to limit the infection. The adaptive immune response develops slowly and needs two weeks to generate the most specific and effective defensive tools. However, the vast majority of PAD patients infected with SARS-CoV-2 did not show signs of hyper-activation of the innate immunity [3-5], an effect possibly due to the immunomodulatory effects on innate immune cells due to the regular use of polyvalent immunoglobulins [30], and a poor adaptive immunity. This pattern of immune responses resembles what we have already shown in asymptomatic immunocompetent subjects [36], and further demonstrated that a balanced cytokine production resulting from a functional but not hyper-reactive innate immunity and a poor adaptive immunity are the conditions associated with an early benign COVID-19 course. Virus-neutralizing antibodies are only one of the correlates of protection for COVID-19, as we and others [4-7] demonstrated in patients with PAD, which, while unable to produce any antibodies, might remain asymptomatic after contracting SARS-CoV-2 infection.

Our data are partially in contrast to observation reported in small cohorts of PAD, showing that CVID patients are able to respond to the BNT162b2 vaccine [31]. Differently from previous studies, we described a homogeneous cohort of patients followed in a single clinical center, analyzing separately CVID and XLA and analyzing different time point for each participant in PAD and in HD group.

In summary, a minority of PAD patients showed adaptive, non-canonical immune responses after SARS-CoV-2 immunization. If these responses to vaccination might result in a partial protection from infection or re-infection is now unknown, since we do not know the levels of antibodies or the frequency of specific B- and T-cells required to protect from the infection. It should be remembered here that each PAD patient should be studied as a unique, in terms of cellular and humoral responses due to the variability of their underlying immune deficiency. In our series, antibody response after two doses of BNT162b2 immunization - overlapping that of HD - was found in one patient homozygous for TNFRSF13B mutation, but not in two patients with a heterozygous TNFRSF13B mutation. In a previous study, we demonstrated that CVID patients with biallelic TNFRSF13B mutations responded also to polysaccharide vaccines, while CVID with only one TNFRSF13B mutation showed an impaired response to vaccination [37].

A major limitation of our study is the short time of observation after vaccination. We do not know if the antibody and cellular responses might persist or decline over time, nor if PAD patients might show delayed responses. However, we do not expect to observe major changes in the immune response to SARS-CoV-2 with time after immunization. It is difficult to hypothesize an improvement of specific B-cell immunity. On the contrary, data available from the protective T-cell immunity after influenza virus vaccination in PAD [11] might suggest a possible strategy aimed to booster the SARS-CoV-2 T-cell specific responses, now poor because induced by a novel antigen, by additional vaccine doses. Since antibody titers are not a surrogate indicator of the magnitude of memory cells [38] our strategy is to follow up our cohort by serological and cellular investigations. The only epidemiological observation we have for now is that one CVID patient experienced an infection by SARS-CoV-2 three months after completing the two-doses vaccination. He remained asymptomatic possibly due to the prompt administration of monoclonal antibodies [39]. For now, our patients might benefit from these new treatments, and possibly from the presence of SARS-CoV-2 antibodies in the coming lots of gammaglobulins [40], regularly used to substitute the missing or partial response to infections and vaccination.

## Data Availability

All data are available in the main text or the supplementary materials.

## Authorship Contributions

AFS, SZ, CFP, FL, RC, and IQ, designed the study and performed data analysis and manuscript preparation. EFS, EPM, ST CQ, MG, CA and SDC performed data experiment. EFS, EPM, ST and FP performed data analysis and helped with manuscript preparation. CM, LB, CA, SA, LR, GC, and GS helped with collection of study samples and clinical information and with manuscript preparation. All authors contributed to the article and approved the submitted version.

