## Supplemental data for "SARS-CoV-2 Vaccine Induced Atypical Immune Responses in Antibody Defects: everybody does their best"

### I. Supplementary Methods

**Cell isolation and cryopreservation.** Heparinized peripheral blood mononuclear cells (PBMCs) were isolated by Ficoll Paque™ Plus 206 (Amersham PharmaciaBiotech) density-gradient centrifugation and immediately frozen and stored in liquid nitrogen until use. The freezing medium contained 90% Fetal Bovine Serum (FBS) and 10% DMSO.

**ELISA for specific IgG, IgA and IgM detection**. A semi-quantitative *in vitro* determination of human IgG and IgA antibodies against the SARS-CoV-2 was performed on serum samples by using the Anti-SARS-CoV-2 Spike ELISA (Euroimmun), according to the manufacturer's instructions. Values were then normalized for comparison with a calibrator. Results were evaluated by calculating the ratio between the extinction of samples and the extinction of the calibrator. The ratio interpretation was as follows: <0.8 = negative, ≥0.8 to <1.1 = borderline, ≥1.1 = positive. To detect IgM anti-RBD we developed an in-house ELISA. 96-well plates (Corning) were coated for 1 h at 37°C with 1 µg/mL of purified SARS-CoV-2 RBD protein (Sino Biological). After washing with PBS 1×/0.05% Tween and blocking with PBS 1×/1% BSA, plates were incubated for 1 h at 37°C with diluted sera (1:100). After washing again, plates were incubated for 1 h at 37°C with peroxidase-conjugated goat anti-human IgM antibody (Jacksons ImmunoResearch Laboratories). The assay was developed with o-phenylen-diamine tablets (Sigma-Aldrich) as a chromogen substrate. Absorbance at 450 nm was measured, and IgM concentrations were calculated by interpolation from the standard curve based on serial dilutions of monoclonal human IgM antibody against SARS-CoV-2 Spike-RBD (Invivogen).

**Detection of antigen-specific B-cells.** To detect SARS-CoV-2 specific B-cells, biotinylated protein antigens were individually multimerized with fluorescently labelled streptavidin at 4°C for one hour. Biotinylated Trimeric SARS-CoV-2 Spike (S1) were purchased from R&D systems. RBD were generated in-house and biotinylation was performed using EZ-LinkTM Sulfo-NHS-LC-Biotin reaction kit (ThermoScientific) following the manufacturer's standard protocol and dialyzed overnight against PBS. Biotinylated Trimeric

Spike was mixed with streptavidin BUV395 (BD) and streptavidin PE (BD) at 25:1 ratio and 20:1 ratio respectively. Biotinylated RBD (gently provided by Takis) was mixed with streptavidin FITC (BD) at 2.5:1 ratio. Streptavidin PE-Cy7 (BD) was used as a decoy probe to gate out SARS-CoV-2 non-specific streptavidin-binding B-cells. The antigen probes prepared individually as above were then mixed in Brilliant Buffer (BD). ~5x$10^6$ previously frozen PBMC samples were prepared and stained with 85μL antigen probe cocktail containing 100ng Spike per probe (total 200ng), 27.5ng RBD and 20ng streptavidin-PE-Cy7 at 4°C for 30 min to ensure maximal staining quality before surface staining with antibodies (listed in table Antibody for staining) was performed in Brilliant Buffer at 4°C for 30 min. MBCs were defined as CD19+CD24+CD27+CD38-Spike+ (S+) or CD19+CD24+CD27+CD38- Spike++ (S++). SARS-CoV-2 specific ATMs were identified as CD19+CD27-CD24-CD38-Spike+ or Spike++; activated MBCs were gated as CD19+CD27+CD24-CD38-Spike+ or Spike++ and specific PBs were identified as CD19+CD24-CD38++CD27++ Spike+ or Spike ++. Stained PBMC samples were acquired on FACS LSRFortessa (BD). At least 4x$10^6$ cells were acquired and analyzed using FlowJo10.7.1 (BD). Phenotype analysis of antigen-specific B-cells was performed only in subjects with at least 10 cells detected in the respective antigen-specific MBCs gate.

**Ex vivo ELISpot Assay for IFNγ detection**

We used an IFNγ ELISpot assay (Mabtech), as described previously (40). Briefly, isolated PBMCs were plated in duplicate, $2\times10^5$ cells/well, stimulated with 1 μg/ml CRUDE PepMix™ SARS-CoV-2 (Spike Glycoprotein, JPT) and incubated for 24 h at 37°C. As a positive control, PBMCs were stimulated with 5 μg/ml of phytohemoagglutinin-P (PHA, Sigma). As negative control PBMCs were plated in serum free CellGenixTM GMP (Cell Genix GMBH). The IFNγ+ spot-forming Unit (SFU) were counted with EliScan (Epson) by Automated ELisa-Spot Assay Video Analysis Systems (A.EL.VIS). Data was presented as the percentage of IFNγ SFUs obtained after pepMix stimulation, respect to the total SFU obtained in the positive control condition (PHA).

**Antibodies for staining**

| | Clone | Catalog number |
|---|---|---|
| CD19 BV786 | SJ25C1 | 563325 |
| CD24 BV711 | ML5 | 563401 |
| CD27 BV510 | T-271 | 740167 |
| CD38 BV421 | HIT2 | 562444 |
| IgG BV650 | G18-145 | 740596 |
| IgM APC | Polyclonal | 709-136-073 |
| Streptavidin PE | | 554061 |
| Streptavidin BUV395 | | 564176 |
| Streptavidin FITC | | 554060 |
| Streptavidin PE-Cy7 | | 557598 |

### II Supplementary Tables

**Table S1.** Demographic, clinical and immunological characteristics of immunized CVID and XLA patients

| **Patients characteristics** | **CVID, immunized n=41** | **XLA, immunized n=6** |
|---|---|---|
| Age (years), median (IQR) | 49 (42-82) | 41 (29-64) |
| Female, n (%) | 26 (59) | 0 (0) |
| Age at PAD diagnosis (years), median (IQR) | 37 (33-72) | 23 (6-33) |
| IgG at diagnosis (mg/dL), median (IQR) | 350 (280-450) | 0 (0-101) |
| IgM at diagnosis (mg/dL), median (IQR) | 22 (7-87) | 0 (0-18) |
| IgA at diagnosis (mg/dL), median (IQR) | 23 (7-181) | 0 (0-24) |
| Chronic Lung Disease, n (%) | 9 (22) | 3 (50) |
| Autoimmunity, n (%) | 16 (41) | 0 (0) |
| Infection, rate per year, median (IQR) | 3 (2-15) | 2 (1-4) |
| CD19+ (%), median (IQR) | 7 (4-30) | 0 |
| CD27+IgM-IgD (%), median (IQR) | 3 (1-40) | 0 |
| CD21low (%), median (IQR) | 32 (6-62) | 0 |
| CD3+ (%), median (IQR) | 73 (68-91) | 76 (70-95) |
| CD3+CD4+ (%), median (IQR) | 35 (29-76) | 42 (39-49) |

**Table S2.** Individual characteristics at the enrollment of 41 CVID and 6 XLA included in the analysis.

| ID | Diagnosis | Age range (years) | Sex | IgG at diagnosis (mg/dL) | IgM at diagnosis (mg/dL) | IgA at diagnosis (mg/dL) | CLD | Autoimmunity | Infection, rate per year | LRTI, rate per year | CD19+ (%) | CD27+IgM-IgD- (%) | CD21low (%) |
|---|---|---|---|---|---|---|---|---|---|---|---|---|---|
| 1 | CVID | 41-45 | F | 320 | 68 | 303 | No | No | 1 | 0 | 5 | 3 | ND |
| 2 | CVID | 56-60 | F | 280 | 2 | 0 | No | No | 5 | 4 | 10 | 0 | 16 |
| 3 | CVID | 76-80 | F | 134 | 13 | 34 | Yes | No | 15 | 0 | 7 | 4 | 46 |
| 4 | CVID | 81-85 | F | 350 | 2 | 75 | Yes | Yes | 5 | 5 | 16 | 5 | 43 |
| 5 | CVID | 56-60 | M | 320 | 7 | 23 | No | No | 2 | 0 | 4 | 2 | 5 |
| 6 | CVID | 71-75 | M | 136 | 0 | 0 | Yes | Yes | 4 | 5 | 17 | 0 | 62 |
| 7 | CVID | 46-50 | F | 301 | 11 | 0 | No | Yes | 12 | 0 | 2 | 5 | 41 |
| 8 | CVID | 46-50 | F | 28 | 3 | 13 | No | Yes | 0 | 0 | 5 | 1 | 2 |
| 9 | CVID | 61-65 | F | 375 | 30 | 181 | No | Yes | 2 | 1 | 13 | 1 | 30 |
| 10 | CVID | 61-65 | F | 312 | 87 | 92 | Yes | No | 0 | 0 | 4 | 21 | 6 |
| 11 | CVID | 46-50 | F | 396 | 25 | 42 | No | No | 2 | 1 | 2 | 7 | 33 |
| 12 | CVID | 51-55 | F | 144 | 20 | 18 | No | Yes | 2 | 0 | 12 | 5 | 46 |
| 13 | CVID | 31-35 | F | 280 | 0 | 0 | No | Yes | 6 | 0 | 10 | 0 | 16 |
| 14 | CVID | 46-50 | F | 105 | 9 | 7 | No | No | 1 | 1 | 10,8 | 2,3 | ND |
| 15 | CVID | 41-45 | M | 269 | 44 | 70 | No | No | 6 | 0 | 6 | 2 | ND |
| 16 | CVID | 56-60 | F | 340 | 7 | 5 | Yes | Yes | 12 | 4 | 12 | 3 | ND |
| 17 | CVID | 21-25 | F | 300 | 0 | 0 | No | Yes | 3 | 0 | 7 | 1 | ND |

| | | | | | | | | | | | | | |
|---|---|---|---|---|---|---|---|---|---|---|---|---|---|
| 18 | CVID | 56-60 | F | 350 | 29 | 66 | No | No | 1 | 1 | 30 | 1 | ND |
| 19 | CVID | 51-55 | M | 380 | 7 | 5 | No | Yes | 7 | 2 | 2 | 10 | 9 |
| 20 | CVID | 26-30 | M | 355 | 53 | 8 | No | No | 2 | 0 | 5 | 7 | ND |
| 21 | CVID | 76-80 | M | 313 | 16 | 52 | Yes | No | 2 | 1 | 15 | 4 | ND |
| 22 | CVID | 31-35 | M | 400 | 6 | 20 | No | Yes | 2 | 0 | 8 | 4 | 9 |
| 23 | CVID | 71-75 | F | 380 | 25 | 52 | No | No | 0 | 0 | 10 | 2 | nd |
| 24 | CVID | 56-60 | M | 360 | 25 | 50 | No | No | 11 | 4 | 8 | 1 | nd |
| 25 | CVID | 66-70 | F | 412 | 55 | 70 | No | Yes | 12 | 5 | 7 | 4 | nd |
| 26 | CVID | 56-60 | F | 350 | 36 | 7 | No | Yes | 6 | 5 | 27,5 | 40 | nd |
| 27 | CVID | 56-60 | M | 253 | 29 | 35 | Yes | No | 10 | 5 | 6 | 5 | 35 |
| 28 | CVID | 26-30 | F | 310 | 0 | 0 | No | Yes | 3 | 2 | 12 | 6 | 40 |
| 29 | CVID | 46-50 | M | 305 | 9 | 8 | Yes | No | 4 | 2 | 4 | 2 | 5 |
| 30 | CVID | 21-25 | M | 387 | 47 | 60 | No | No | 4 | 2 | 15 | 1 | 45 |
| 31 | CVID | 51-55 | F | 280 | 10 | 15 | No | Yes | 5 | 3 | 2 | 4 | 38 |
| 32 | CVID | 21-25 | F | 280 | 20 | 0 | No | No | 1 | 1 | 5 | 1 | 2 |
| 33 | CVID | 26-30 | F | 358 | 39 | 78 | No | Yes | 4 | 3 | 12 | 1 | 28 |
| 34 | CVID | 36-40 | F | 387 | 8 | 15 | No | No | 7 | 4 | 4 | 18 | 4 |
| 35 | CVID | 71-75 | M | 380 | 59 | 18 | Yes | No | 3 | 3 | 2 | 6 | 32 |
| 36 | CVID | 46-50 | M | 450 | 17 | 45 | No | Yes | 3 | 1 | 3 | 1 | 4 |
| 37 | CVID | 41-45 | F | 432 | 7 | 25 | No | No | 2 | 1 | 18 | 0 | 55 |
| 38 | CVID | 26-30 | M | 400 | 30 | 40 | No | Yes | 2 | 1 | 2 | 4 | 40 |

| | | | | | | | | | | | | | |
|---|---|---|---|---|---|---|---|---|---|---|---|---|---|
| 39 | CVID | 46-50 | M | 412 | 25 | 55 | No | No | 3 | 1 | 4 | 1 | 2 |
| 40 | CVID | 56-60 | F | 400 | 50 | 75 | No | No | 2 | 0 | 12 | 1 | 32 |
| 41 | CVID | 41-45 | F | 358 | 23 | 5 | no | No | 0 | 0 | 3 | 4 | 45 |
| 42 | XLA | 26-30 | M | 0 | 0 | 0 | No | No | 3 | 2 | 0 | 0 | 0 |
| 43 | XLA | 61-65 | M | 0 | 0 | 0 | Yes | No | 1 | 1 | 0 | 0 | 0 |
| 44 | XLA | 46-50 | M | 0 | 0 | 0 | Yes | No | 0 | 0 | 0 | 0 | 0 |
| 45 | XLA | 31-35 | M | 101 | 0 | 0 | No | No | 0 | 0 | 0 | 0 | 0 |
| 46 | XLA | 51-55 | M | 30 | 18 | 24 | Yes | No | 4 | 4 | 0 | 0 | 0 |
| 47 | XLA | 21-25 | M | 0 | 2 | 3 | No | No | 3 | 1 | 0 | 0 | 0 |

Abbreviation: M: male, F: Female; CLD: Chronic Lung Disease; LRTI: Low Respiratory Tract Infections,

### II. Supplementary Figures

**Figure S1.** Gating strategy to identify RBD+ cells inside total Spike positive (S+ plus S++) MBCs. Flow cytometry plots in one healthy control and one CVID patient showing the staining pattern of RBD+ MBCs. In the CVID patient RBD+ MBCs are undetectable.

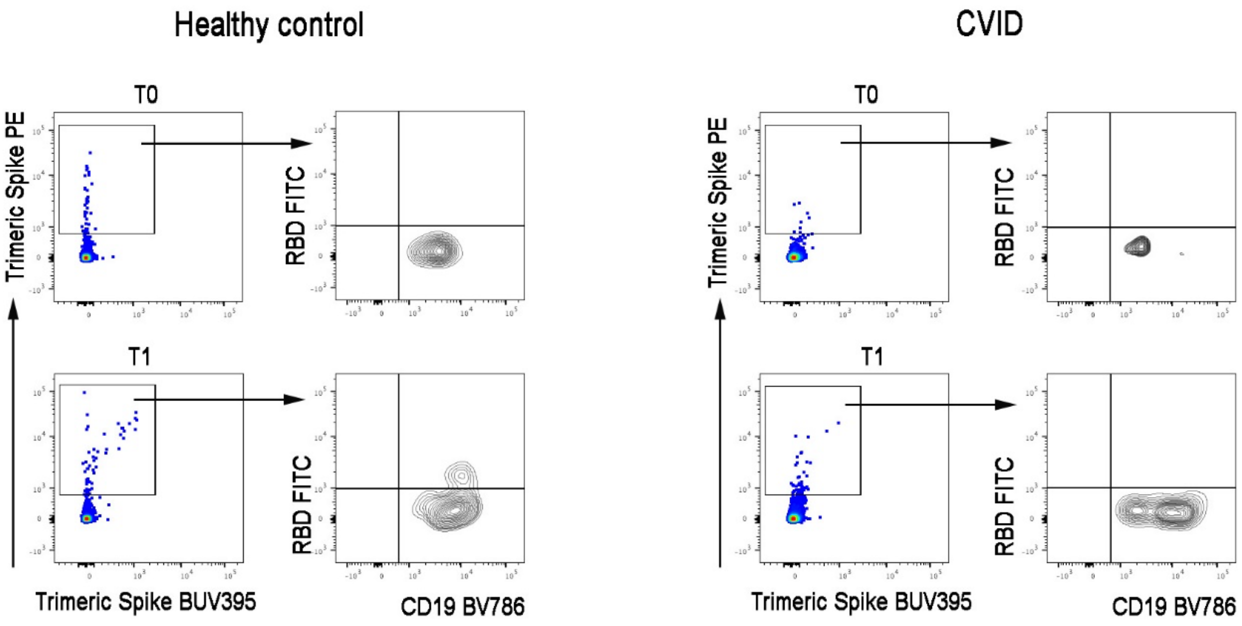
